# MAJIQ-CLIN: A novel tool for the identification of Mendelian disease-causing variants from RNA-Seq data

**DOI:** 10.1101/2025.01.30.25321185

**Authors:** Joseph K Aicher, Dina Issakova, Barry Slaff, San Jewell, Nicholas F Lahens, Gregory R Grant, Diana Baralle, Jill A Rosenfeld, Daryl A Scott, Undiagnosed Diseases Network, Elizabeth J Bhoj, Yoseph Barash

**Affiliations:** Department of Genetics, Perelman School of Medicine, University of Pennsylvania (Philadelphia, USA); Department of Biology, School of Arts and Sciences, University of Pennsylvania (Philadelphia, USA); Department of Computer and Information Sciences, School of Engineering, University of Pennsylvania (Philadelphia, USA); Faculty of Medicine, University of Southampton (Southampton, UK); Children’s Hospital of Philadelphia (Philadelphia, USA); Baylor Genetics Site, Undiagnosed Diseases Network

## Abstract

The current diagnostic rate for patients with suspected Mendelian genetic disorders is only 25 to 58%, even though whole exome sequencing (WES) is part of the standard of care. One reason for the low diagnostic rate is that traditional WES analysis methods struggle to detect RNA splicing aberrations. It is estimated that 15-50% of human pathogenic variants alter splicing, with numerous splice-altering variants being causal for known Mendelian disorders. Developing reliable diagnostic tools to detect, quantify, prioritize, and visualize RNA splicing aberrations from patient RNA sequencing is therefore crucial. We present MAJIQ-CLIN, a method to address this need to augment clinical diagnostic using RNA-Seq and compare it to existing tools. We include the first systematic evaluation of the accuracy of such tools using synthetic data across several aberration types and transcript inclusion levels; we also evaluate accuracy on several datasets of biologically validated solved test cases. We show that MAJIQ-CLIN compares favorably to existing tools in both accuracy and efficiency, then use MAJIQ-CLIN to investigate several unsolved patient cases from the Undiagnosed Diseases Network.

## Introduction

Over 400 million people worldwide suffer from over 7000 rare diseases [1]. Approximately 80% of all rare disease cases are genetic in origin, and most are Mendelian [2] [3]. Whole Exome Sequencing (WES) is the state-of-the-art genetic test for diagnosing patients with suspected Mendelian disorders. Unfortunately, the diagnostic rate of WES is only 25 to 58% [4] [5]. Whole Genome Sequencing (WGS) is able to improve that rate by another 34% [6], yet millions of patients still lack diagnosis.

RNA-Seq is a promising diagnostic tool because it directly detects pathological changes in a patient’s transcriptome. These pathological changes include intronic or synonymous mutations, often overlooked by DNA sequencing methods. Recent works show that significant improvement can be achieved by integrating RNA-Seq into existing WES or WGS pipelines: Deelen et al. [7] identify likely causal genes for ten patients undiagnosed after WES using RNA-Seq, and Deshwar et al. [8] and Riquin et al. [9] show how RNA-Seq can help interpret the impact of variants in patients. In particular, a key process missed by WES/WGS is aberrant alternative RNA splicing (AS), where ‘incorrect’ exonic or intronic segments of a pre-mRNA are spliced together to form the mRNA. Errors in alternative splicing can lead to non-functional isoforms, early termination codons, or frameshift mutations, resulting in nonsense-mediated decay and RNA degradation. Over 90% of the human genes naturally undergo AS [10], yet approximately 35% of pathogenic variants alter splicing [11]. Thus, detecting these deleterious variants is crucial for comprehensive genetic diagnostics.

Detecting splicing aberrations for the purpose of clinical diagnostics presents several challenges. First, it may not be immediately clear which clinically accessible tissue should be sequenced. An accessible tissue (e.g., blood) may not capture a patient’s expression or splicing aberrations for a tissue of interest (e.g., brain), although tools do exist to detect which accessible tissue is best for a particular case [12]. Still, the challenge of efficiently and accurately detecting potentially causal splicing aberrations in the patient persists. This challenge is corroborated by the fact that AS is a noisy process. Varying degrees of natural variability have been observed across individuals [13], and additional variability due to technical factors such as batches is inevitable [14].

Several previous works have proposed tools to apply RNA-Seq to clinical diagnostics. Cummings et al. [6] filtered patient RNA-Seq for splice junctions not found in controls, successfully diagnosing 17 patients. Gonorazky et al ([15]) expanded upon this approach, allowing candidate splice junctions to be present in a few samples in their control dataset. Later works looked for outlier splice variations across all genes using different models. LeafCutterMD [16] provided a statistical framework that improves upon the previously published LeafCutter [17] in detecting outlier splicing events. FRASER [18] is another alternative approach, recently releasing a second iteration (FRASER2) [19]. These approaches served as a substantial proof of concept and helped solve several clinical cases [20][21], but a systematic comparative evaluation of analysis methods has not been performed.

To allow us to evaluate these tools, we defined several requirements that must be met by a tool suitable for clinical diagnostics. First, it must accurately detect and quantify patients’ splicing aberrations, including unannotated and complex variants. Variants where splicing differs significantly between the patient and a set of healthy controls should be prioritized into a list for the clinician to inspect, with true causal variants highly ranked. It should also be able to correct for known and unknown confounders (eg. batches) to minimize false positives. The entire process should be highly efficient in time and memory, avoiding unnecessary re-processing of data (e.g., control cohorts) when new patients are added. Finally, the tool should include a user-friendly visualization to simplify the complexity of splicing variations.

We evaluated FRASER and LeafCutterMD on these criteria and also developed our own tool, MAJIQ-CLIN. MAJIQ-CLIN is based on an updated (V3) version of the MAJIQ software [22] [13] [23] for RNA splicing detection, quantification, and visualization. MAJIQ-CLIN is able to capture non-annotated and/or patient-specific splicing events, formulated as Local Splicing Variations (LSVs). LSVs are defined as a set of splice junctions coming either into or from a single reference exon and a collection of LSVs for a gene forms a splicegraph. As illustrated in Figure 1A, the LSV formulation allows MAJIQ-CLIN to detect and quantify not only ‘classical’ AS events such as cassette exons but also complex splicing events involving more than one alternative RNA segment. MAJIQ quantifies the inclusion of each junction in an LSV as Percent Spliced In (PSI, or Ψ). Since Ψ is not directly observable, MAJIQ uses the observed read count and positions to estimate a posterior distribution of possible Ψ values for a junction (see Methods). To detect outliers, MAJIQ-CLIN assesses the gap between extreme quantiles of the Ψ distributions for the controls and an individual patient (Figure 1B), called the Ψ-GAP. MAJIQ-CLIN then uses the Ψ-GAP to call two categories of junctions as outliers. The first are junctions that are unique or almost unique to the patient, termed Private LSVs (pLSVs), hence their inclusion level in controls is zero. The second category is those junctions that are observed in controls but with a large Ψ-GAP, termed Outlier LSVs or oLSVs. MAJIQ-CLIN then ranks and prioritizes these variants for the clinician based on their uniqueness to the patient and the Ψ-GAP. MAJIQ-CLIN also incorporates MOCCASIN [14] to automatically detect and correct for known and unknown confounders between samples, addressing within-sample covariation. Finally, CLIN adds new components to the visualization package VOILA so users can easily inspect and analyze the ranked pLSVs and oLSVs.

**Figure 1:**
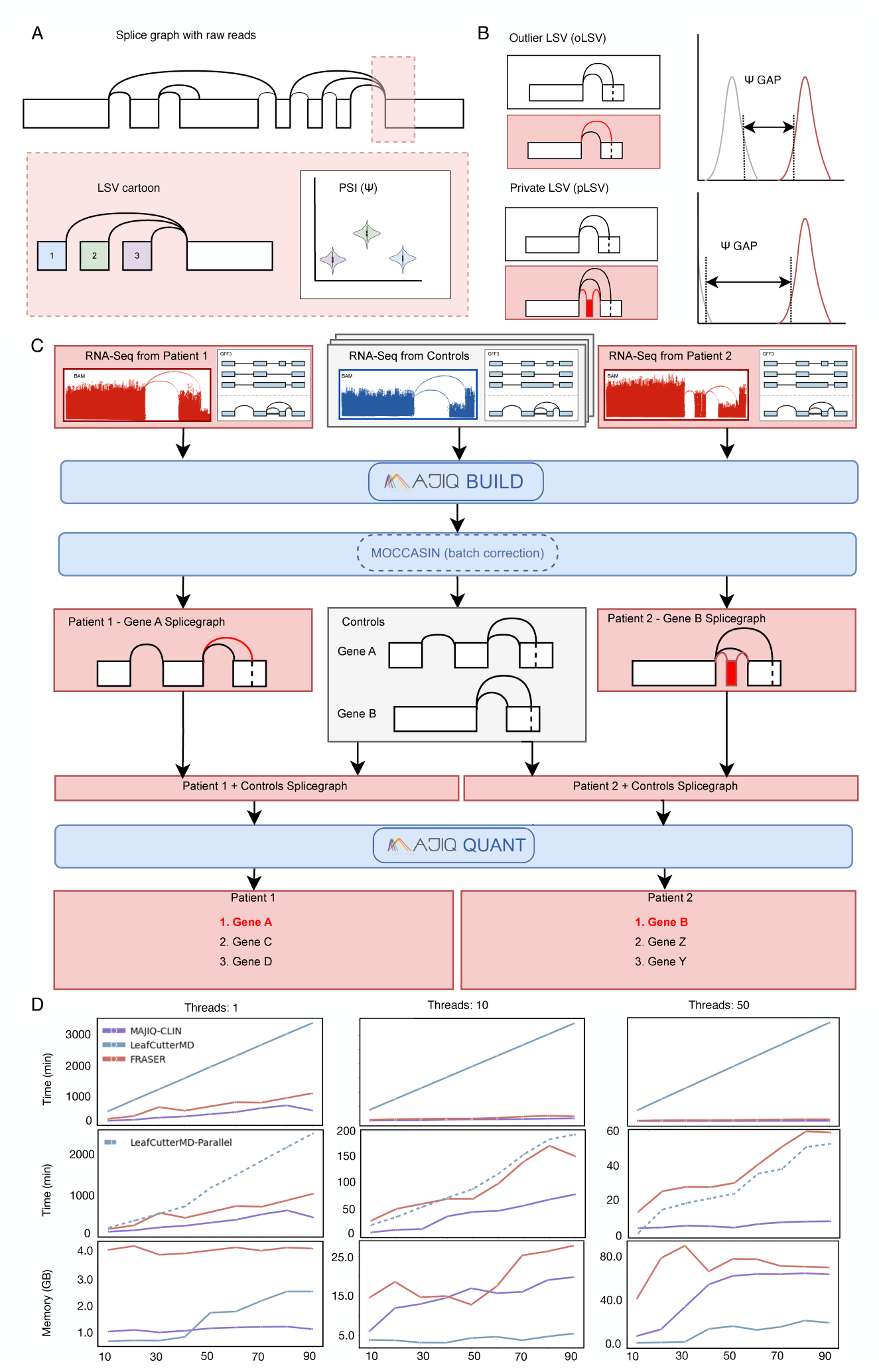
**A**: MAJIQ quantifies splicing using splicegraphs, made up of Local Splicing Variations (LSV). An LSV is defined as a set of junctions coming into or out of a reference exon. For each junction, MAJIQ estimates Percent Spliced In (PSI, or Ψ), a measure of junction usage. **B**: MAJIQ-CLIN detects two types of outliers, outlier LSVs (oLSVs) and private LSVs (pLSVs). oLSVs are outliers where the splicegraph between the patient and controls is the same, but PSI is different. pLSVs are splicing variants unique to the patient, contained in at most `min-experiments` (user-defined, default 1) cases in the control set.**C**: MAJIQ-CLIN takes as input RNA-Seq data from patients and controls, as well as a GFF3 annotation. Then, MAJIQ-BUILD builds a splicegraph for each gene and performs confounder correction. pLSVs are detected by comparing patient and control splicegraphs. CLIN then creates combined splicegraphs for each patient and the control set, quantifying LSVs using MAJIQ-QUANT. CLIN then outputs a list of candidate LSVs and genes to the clinician, ordered by type (pLSV, oLSV) and Ψ-GAP. **D**: Runtime and memory usage comparison with 1, 10, 50 threads. Top row: LeafCutterMD run with the default bam-to-junc step (unparallelized). Middle row: Same as above but with in-house script added to parallelize LeafCutterMD bam-to-junc (dashed line).

Getting a precise assessment of how well MAJIQ-CLIN or other tools detect different types of splicing events is challenging, since most applicable datasets are small. For this purpose, we created the first of its kind large-scale “realistic” synthetic dataset to compare tools with respect to the criteria described above. This dataset includes synthetic RNA-Seq samples based on GTEX, with clean “control” samples and “patient” samples with artificially introduced splicing aberrations (both pLSVs and oLSVs). Using this data, we compared MAJIQ-CLIN, FRASER2, and LeafCutterMD. To the best of our knowledge, this analysis is the first to assess the ability of such tools to detect splicing aberrations as a function of the magnitude of change and the type of splicing variation (e.g. exon skipping or intron retention).

In the following results, we first introduce MAJIQ-CLIN and describe its efficient use of resources. We then compare its performance to that of its competitors with the aforementioned synthetic dataset, demonstrate its performance on real datasets of solved biological cases, and discuss the effect of confounder correction and choice of control cohort. We find that FRASER and LeafCutterMD fulfill some, but not all, of the requirements for a clinical RNA-Seq splicing tool, with MAJIQ-CLIN filling that gap. Finally, we apply MAJIQ-CLIN to unsolved cases from the Undiagnosed Diseases Network [20], showing its promise as a clinical diagnostic tool.

## Results

### 0.1 The MAJIQ-CLIN pipeline offers efficient integration of patient and control samples

The MAJIQ-CLIN workflow is illustrated in Figure 1C. The pipeline takes as input BAM files from patients, healthy controls, and a genome annotation in GFF3 format. The controls are processed as a group by MAJIQ-BUILD, which outputs a set of LSVs as a splicegraph. Each patient sample is processed individually in the same way so that samples can be added incrementally without re-processing previous samples. Optionally, the pipeline can combine the LSVs across the controls and patients to correct confounding factors using MOCCASIN [14]. MAJIQ-CLIN then builds the union set of LSVs between the controls and each patient and applies MAJIQ-QUANT to quantify the read rate for each junction. Junctions’ read rate are then used to calculate posterior probabilities for Percent Spliced In (PSI, or Ψ), an estimate of junction inclusion. Consequently, MAJIQ-CLIN detects and reports two kinds of potentially important LSVs: Private LSVs (pLSVs) that are present in the patient but are very rare or nonexistent in the control population, and Outlier LSVs (oLSVs) where CLIN detects junctions that are present in both the controls and the patient but with significantly different inclusion levels. MAJIQ-CLIN then prioritizes the detected pLSVs and oLSVs by their Ψ-GAP, but pLSVs are ranked first by default as they are more likely to be clinically significant. A gene list is output to the clinician and can be visualized using our visualization software [22].

To assess the efficiency of the MAJIQ-CLIN pipeline we analyzed it, along with previously published tools, in terms of runtime and memory consumption. To create a realistic usage scenario we ran groups of 10-90 samples as leave-one-out controls with 1, 10, and 50 threads on a computational cluster. For LeafCutterMD, the runtime for the bamtojunc step was estimated by multiplying the average runtime times the number of samples (see Methods). Figure 1D shows the results. In terms of memory, MAJIQ-CLIN uses slightly more memory than LeafCutterMD for all but the smallest sample sizes in a single-thread run. However, those differences are arguably not significant. Specifically, using one or ten threads, MAJIQ-CLIN’s memory usage remains manageable on an average desktop or even a laptop, without requiring a computational cluster. However, users with access to more memory on a computational cluster can take advantage of 50 threads as shown in the right panel. FRASER required more memory than MAJIQ-CLIN for almost all numbers of samples tested.

While memory usage of CLIN and LeafCutterMD remains similar, MAJIQ-CLIN performs significantly faster than its competitors, thus justifying the slightly higher memory use. While FRASER is still slower than MAJIQ-CLIN, both are dwarfed by LeafCutterMD’s runtime. Most of this runtime was due to the time taken by bamtojunc (estimated; see Methods). We then parallelized the bamtojunc step for LeafCutterMD as suggested in the documentation (1D, row 2). The runtime of this parallelized version remained significantly slower than MAJIQ-CLIN. Thus, for larger sample sizes typical of a clinical diagnostics pipeline, MAJIQ-CLIN delivers substantially faster results and more effectively utilizes provided computational resources than both of its competitors.

### 0.2 MAJIQ-CLIN significantly improves detection of patient specific splicing aberrations

After assessing the efficiency of MAJIQ-CLIN in terms of memory and runtime we turned to perform a comprehensive assessment of its accuracy. We first evaluated detection of rare variants unique to the patient (pLSVs), as these are more likely to be clinically significant [24]. A tool aiming to detect aberrant splicing from patient RNA-Seq must be able to detect and flag these variants across different levels of transcript inclusion and a variety of splicing events, such as exon truncation or extension, exon skipping or addition, and intron retention. To assess pLSV detection we thus created an extensive synthetic dataset with “patients” and “controls.” Briefly, [13] 300 cerebellum and skeletal muscle tissue samples from the Genotype-Tissue Expression (GTEx) Portal served as the basis for 300 matching simulated samples. Each biological sample was quantified by RSEM (which is unrelated to MAJIQ or MAJIQ-CLIN) and then those quantifications were used by the BEERS simulator [25] to create a matching ‘realistic’ synthetic sample. (See Methods). In the same way, four additional samples served as the base for the “patients.” For the patient samples, we generated five types of splicing aberrations not found in the controls: new exons, exon skipping, intron retention, and changes to 3’ or 5’ splice sites. We then randomly sampled from this list to create 100 synthetic “patients,” containing 5-100 aberrant transcripts at varying levels of Ψ from 0.05 to 0.5 (See Methods). We note this setting with limited number of aberrations per sample was chosen to accommodate for the FRASER model. Specifically, since FRASER’s confounder correction is built into its autoencoder including many aberrations in a sample led to aberrant genes filtered out and low performance.

Panels A and B of Figure 2 show the pLSV precision-recall curves based on this synthetic data for MAJIQ-CLIN, FRASER and LeafCutterMD. MAJIQ-CLIN consistently shows higher precision and recall than FRASER and LeafCutterMD at more stringent thresholds, performing reliably across different types of splicing aberrations and levels of variant inclusion (Ψ). LeafCutterMD also struggles with intron retention — a splicing event increasingly recognized for its role in gene expression regulation. Many retained introns contain premature stop codons that trigger nonsense-mediated decay (NMD) pathways [26] [27], but some bypass NMD and have been associated with 16 types of cancer [28] and Alzheimer’s disease [29]. MAJIQ-CLIN demonstrates high precision across this and other events, making it a robust tool for detecting clinically significant splicing variants. Notably, the ability to prioritize pLSVs significantly boosts MAJIQ-CLIN’s performance though even when this flag is removed (dashed line) it still performs well in pLSV detection compared to the other tools.

**Figure 2:**
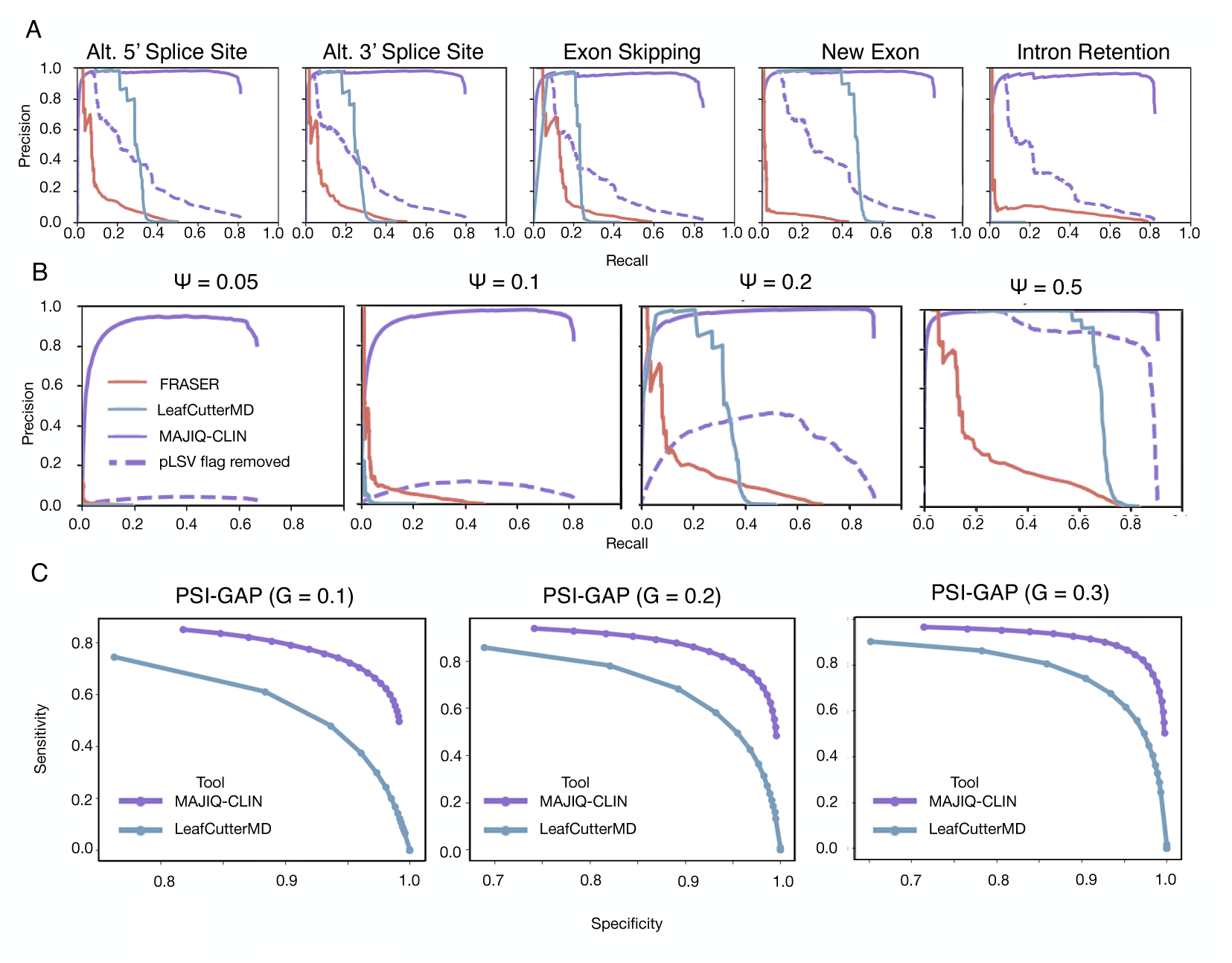
A: Comparison of pLSVs detection in synthetic data across different splicing aberration types. Dotted line shows results when positives are not restricted to pLSVs identified at the splicegraph stage. B: Same data as in A but broken down by levels of variant inclusion (Percent Spliced in, or PSI). C: Comparison of oLSVs detection from synthetic data based on tissue-specific splicing differences from GTEx for different. Three levels of Ψ-GAP were tested (*G* ∈ {0.1, 0.2, 0.3}). FRASER was not included as it failed to run on this data (see Methods).

Next, we turned to evaluate MAJIQ-CLIN and its competitors on variants shared between the patient and the controls but with significantly different junction usage (Ψ) in the patient, or oLSVs. As AS varies across tissues, GTEx already contains oLSVs when comparing samples from different tissues. Thus, we randomly selected 20 cerebellum and 20 skeletal muscle synthetic samples from the dataset described above to serve as the “patients” for this analysis. We then compared each such “patient” to the “controls” from the other tissue - *i.e.*, each skeletal muscle “patient” was compared with the 150 cerebellum “controls.” Then, for each ‘patient’ sample we defined as ground truth labels splicing variations with a known distance larger than a defined threshold Ψ*_GAP_ > G* between the distribution of Ψ estimates for the ‘patient’ and ‘control’ (see Methods). We tested three values for the threshold *G* ∈ {0.1, 0.2, 0.3}, generating precision-recall curves for each. The results of this analysis, shown in panel C of 2, show MAJIQ-CLIN outperforms LeafCutterMD significantly and demonstrates high performance on oLSVs across all *G* thresholds used. FRASER was not included in this evaluation as we observed that including too many splicing variations in a single sample caused FRASER to auto-correct it as an outlier, therefore failing to report the expected splicing aberrations.

### 0.3 Confounder corrections, control set choice and size can significantly affect outlier detection

To make RNA-Seq usable for clinical diagnostics it is important that a tool for detecting aberrant splicing should return a short list of candidate genes, with the true causal variant ranked highly on the list. A tool’s ability to do so depends significantly on the dataset used, particularly on the choice of control dataset. A larger control set improves the ability to detect pLSVs and reduces false positives, while a smaller set may not adequately represent the population. On the other hand, including many control samples that are too dissimilar to the patient sample can introduce confounding factors into the analysis and inflate the number of reported aberrations. Correcting for confounders is therefore important to reduce false positives. To control for potential confounders MAJIQ-CLIN integrates MOCCASIN [14], which can be run with varying number of unknown confounders to detect and correct in the data. We therefore sought to assess how the choice of control set and the number of confounders corrected on may affect real dataset analysis with MAJIQ-CLIN.

First, we assessed the effect of confounder correction and control set definition using 55 whole blood RNA-Seq samples from patients with suspected rare Mendelian disorders [21]. Notably, this data is composed of three batches of varying sizes. We ran MAJIQ-CLIN on the patients in each batch with three choices of controls: the remainder of the same batch, the remainder of the batch and the other batches in the dataset, and a set comprising the remainder of the batch and the other batches augmented with GTEx blood samples. The results are shown in Figure 3A. As expected, we find that the number of identified aberrations depends strongly on the choice of controls. For instance, when the batch is small (n=7) more outliers are reported, especially when using the same small batch as the only control, and adding the other batches to the control set helps shorten the list of reported aberrations. This effect is smaller when the batch is of size 16 but when the batch is over 30 it seems that using only the same batch as a control set results in the shortest splicing aberrations list. In all cases adding samples that are significantly different (GTEx) increases the number of reported aberrations. Conversely, increasing the number of unknown confounders significantly decreases the number of reported aberrations. We note that while the above results were derived from the three different batches in the data similar results are observed when down-sampling the large batch (3B). We also do not suggest using solely external controls without using the remainder of the same batch as a leave-one-out control, as this significantly increased the length of the outlier list.

**Figure 3:**
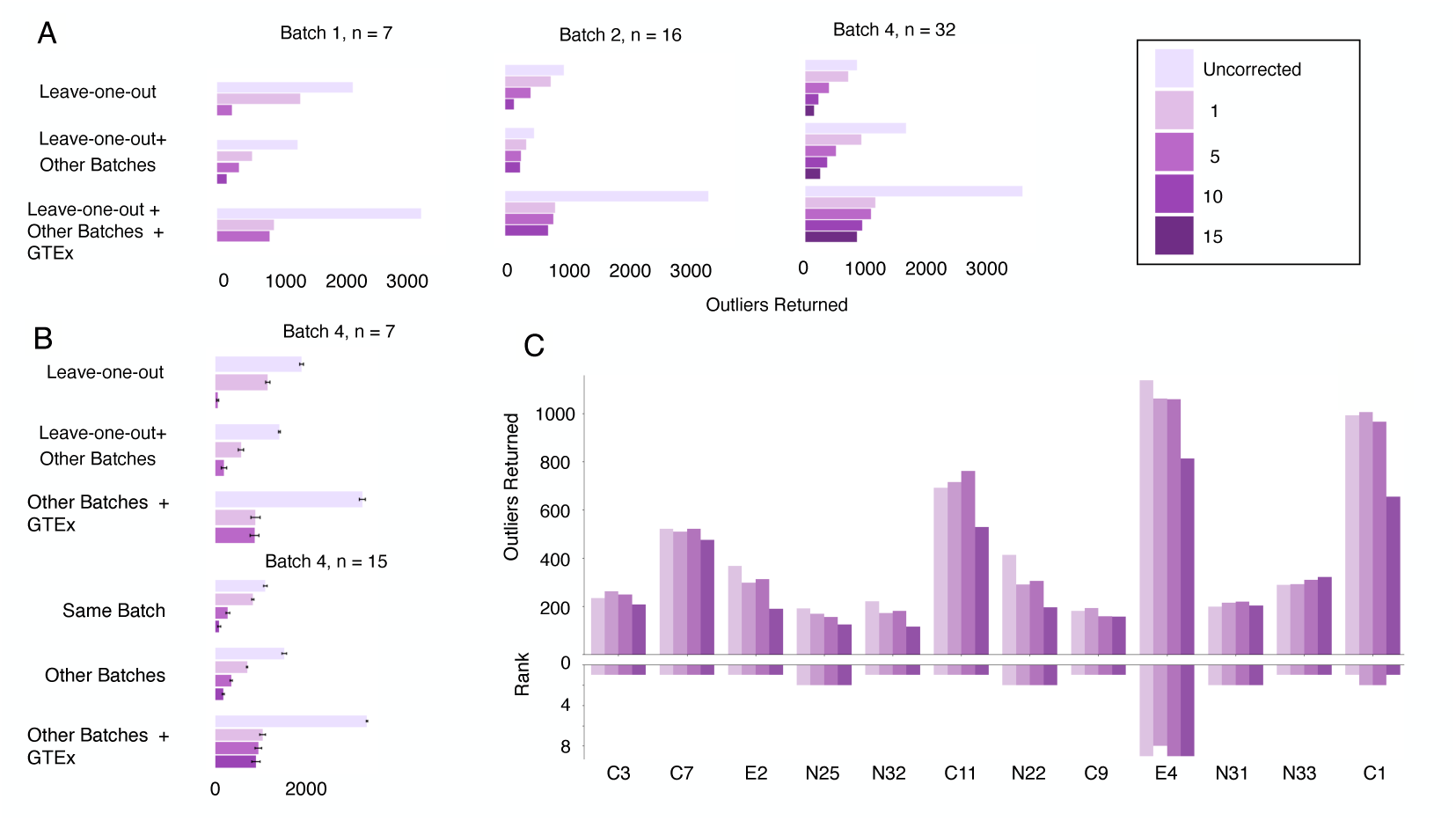
Confounder correction decreases length of list returned to clinician, dependent on batch size and choice of control set. **A:** The total number of outliers identified when running MAJIQ-CLIN on three batches of varying sizes from Baralle et al., with different choices of controls (2024), with different control sets and numbers of unknown confounders corrected. **B:** Sub-sampling from batch 4 exhibits a similar trend, further demonstrating the influence of batch size. **C:** The number of outliers detected when analyzing cases with previously identified splicing variants from Cummings et al. (2017), alongside the ranking of causative genes in the resulting list.

One caveat with the above analysis is that we can not assess the effect on true casual variants as those are generally unknown. To assess effects on casual variants detection and ranking we thus turned to a second dataset from Cummings et al. [6]. This dataset contains RNA-Seq samples from 53 patients, of which 16 are identified splicing variants. For each of these 16 patients, the remaining samples were used for controls. The results are shown in Figure 3C. MAJIQ-CLIN successfully identifies the causal genes in all cases and ranks them close to the top of the list. Similar to the Baralle dataset, the use of MOCCASIN reduces the length of the list returned for many cases (disease-associated outliers are shown for scale), but does not significantly affect the ranking of causal variants.

### 0.4 MAJIQ-CLIN offers improved interpretability and visualization that aid in analyzing patient data

Beyond detection and high ranking of causal splicing aberrations, a tool that aids in clinical diagnostic must support easy to interpret visualization of the aberrations it detects. For this, MAJIQ-CLIN incorporates new features into the VOILA visualization package to allow direct visualization of junctions and LSVs which differ between cases and controls. Specifically, all junctions and LSVs found in the patient or case group (see Methods) but not the controls can be highlighted in the gene’s splicegraph. These aberrations are also indexed and searchable within VOILA. To showcase this new visualization and MAJIQ-CLIN’s utility in a real-life clinical setting, we applied it to undiagnosed patient cases from the Undiagnosed Diseases Network (UDN) [20]. We used fibroblast RNA-Seq data from the patients with other cases as controls. To focus our search we filtered the results to consider only genes associated with disease by OMIM and containing a pLSV variant.

### 0.5 Case studies using MAJIQ-CLIN

Here, we highlight several patient cases where MAJIQ-CLIN identified variants of potential clinical significance missed/not reported in previous publications. These cases were chosen to show different use cases for MAJIQ-CLIN. FRASER was previously used in this set to detect splicing aberrations, leading to the diagnosis of several new cases [20]. We filtered for genes containing pLSVs and associated with a Mendelian disease in OMIM [3], which resulted in lists of up to 2 genes per case.

#### Case 1 - MAJIQ-CLIN finds unknown diagnosis

The first case we highlight involves a male patient age 11-15. The UDN phenotype record includes vision-related problems (astigmatism, myopia), cerebellum ataxia and hypoplasia, delayed speech and language development, gait abnormalities, low muscle tone, and distinctive dysmorphic features. MAJIQ-CLIN finds a pLSV variant in *POLR3B*, a gene responsible for building a subunit of RNA Polymerase III. MAJIQ-CLIN showed that this variant causes the skipping of exon 20 in the reference transcript. As MAJIQ-CLIN cannot distinguish between an alternatively spliced junction with Ψ = 0.5 and a constitutively spliced but heterozygous variant, it was necessary in this case to look at the proband’s parentage. As shown in Figure 5, the proband does indeed have a paternally inherited heterozygous exon 20 deletion. A copy number variant was also found in this gene using optical genome mapping. Recessive mutations in this gene have been associated with Pol-III-related leukodystrophy [3], typically presenting with ataxia, broad unsteady gait, vision problems such as restricted eye movement, nearsightedness, and cataracts, as well as intellectual disability and delayed speech development. These symptoms are consistent with the symptoms reported for this patient by UDN. Although hypomyelination is not explicitly listed as a symptom, the patient exhibits symptoms that could be consistent with hypomyelination, such as cerebellar ataxia and hypoplasia. The patient also lacks a common symptom of hypodontia; however, several documented cases do not present with this symptom [30]. Importantly, upon contacting the UDN researchers about this finding we learned they confirmed the heterozygous exon 20 POLR3B deletion using optical genome mapping and genome sequencing. This deletion was inherited *in trans* with a likely pathogenic POLR3B missense variant from the mother. We note that MAJIQ-CLIN detected this variant independently using RNA-Seq without any DNA-based sequencing. This case demonstrates the value of MAJIQ-CLIN in a clinical setting.

**Figure 4:**
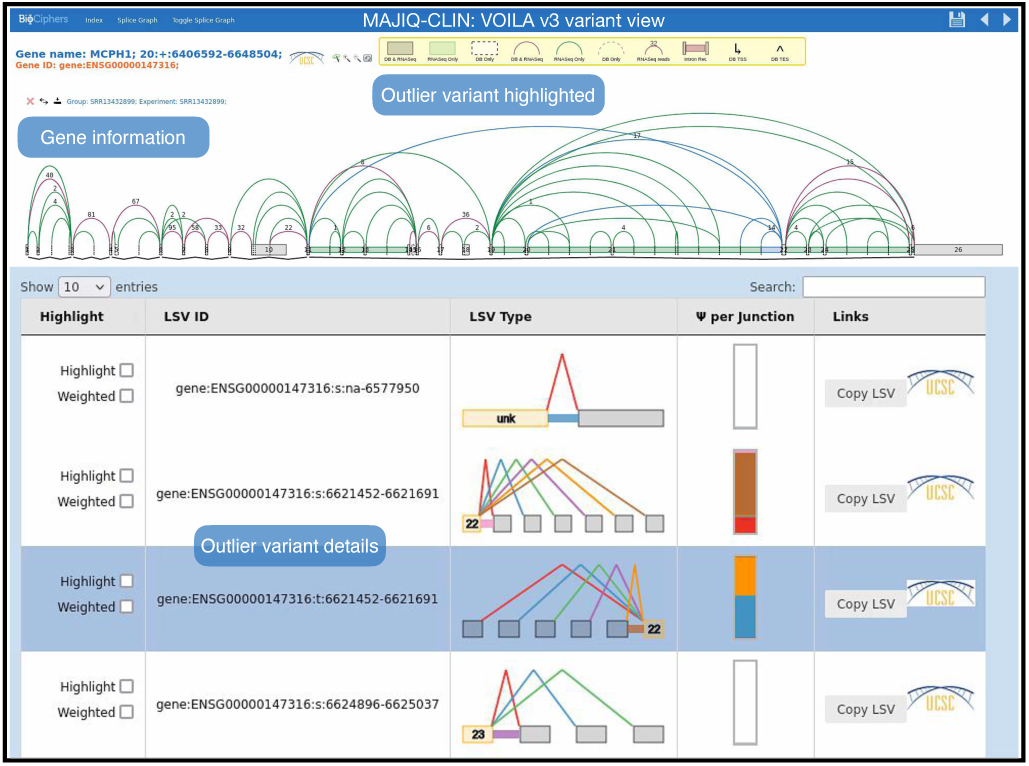
The VOILA visualization of MAJIQ-CLIN results highlights LSVs found in the annotation in gray, those identified exclusively in the RNA-Seq data in green, and pLSVs in blue, which are also emphasized in the individual pLSV view.

**Figure 5:**
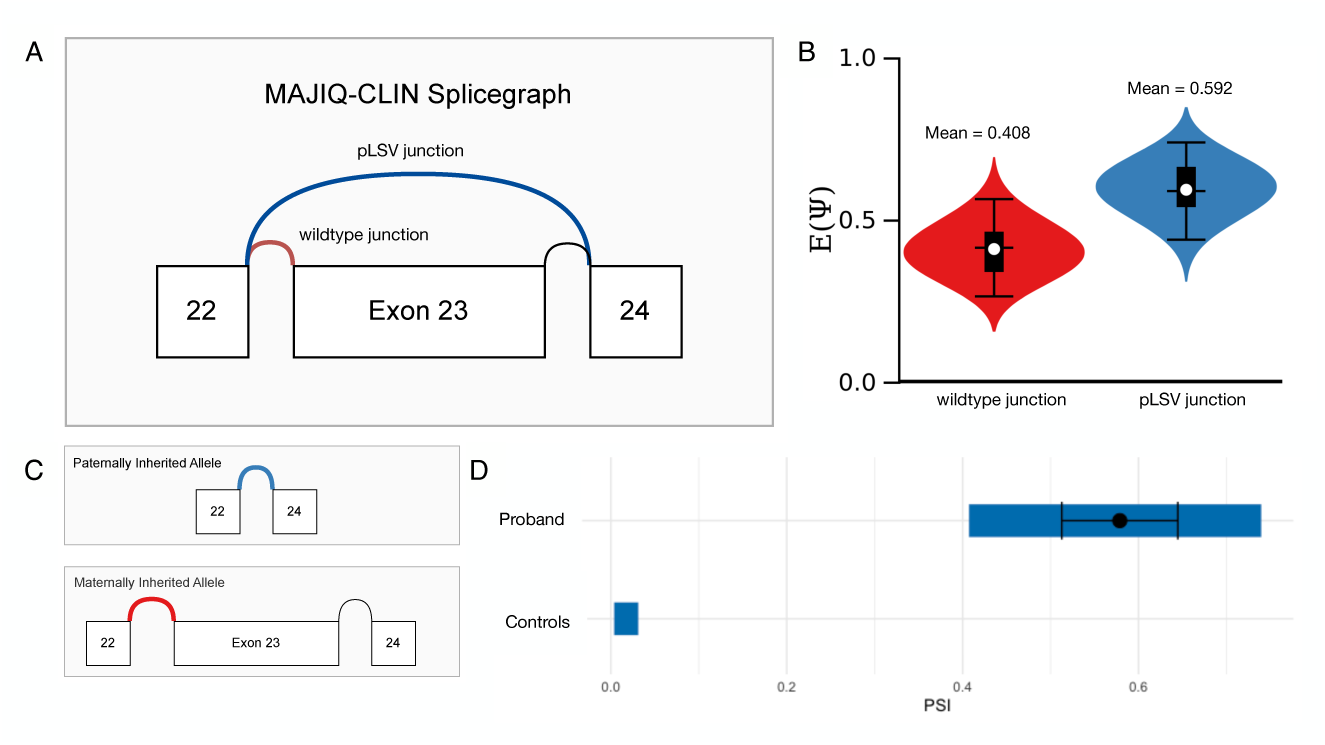
**A**: Splicing variant skipping exon 23 (exon 20 in the reference transcript ENST00000228347) of POLR3B found by MAJIQ-CLIN. **B**: PSI distributions of novel and wildtype junctions in the proband. **C**: Paternally and maternally inherited alleles in the proband as shown by optical genome mapping. **D**: Usage of novel junction in proband compared to controls (shaded area: 5th to 95th percentile; mean and standard deviation shown for proband).

#### Case 2 - MAJIQ-CLIN finds unknown diagnosis

This case involves a female patient with symptom onset at birth. The UDN phenotype record lists a number of symptoms including some physical traits (high palate, pes planus, coxa valga), vision-related issues (esotropia, anomalies of visually evoked potentials, and astigmatism), global developmental delay, and specifically issues related to the acquisition of gross motor development (general delays, broad gait, involuntary movements, and reduced tendon reflexes).

This patient has a complex pLSV variant in the gene MCOLN1. Mutations in MCOLN1 are associated with Mucolipidosis IV, a disease involving severe impairment in the acquisition of psychomotor skills, visual impairment stemming from clouding of the cornea and retinal degeneration, and hypotonia (affecting tendon reflexes). However, as this is an autosomal recessive gene, this diagnosis is less definite than in Case 1 described above.

#### Case 3 - MAJIQ-CLIN finds additional diagnosis

This case involves a female patient age 1-5. She had been diagnosed with Long-chain 3-hydroxyacyl-CoA dehydrogenase deficiency (LCHAD), which was linked to a number of her symptoms, such as heart issues, structural and functional abnormalities, ophthalmic issues, and problems with metabolism. However, this patient also had a number of kidney-related symptoms: nephrotic syndrome, focal segmental glomerulosclerosis, and elevated creatinine levels - unexplained by her LCHAD diagnosis.

MAJIQ-CLIN analysis revealed a complex pLSV variant in the ARGHDIA gene, associated with nephrotic syndrome type 8, suggesting a potential root cause for the nephrosis. This secondary diagnosis would explain the remainder of the patient’s symptoms, and this case shows the potential of MAJIQ-CLIN to resolve finer details of a diagnosis.

#### Case 4 - MAJIQ-CLIN Suggests Novel Disease Mechanism

The patient in this case presents as tall, with an oval face, high forehead, prominent nose, and macrocephaly. Additional phenotypes included myopia, striae distensae, joint hypermobility, mitral valve prolapse, pes planus, and pneumothorax.

MAJIQ-CLIN analysis revealed a splicing aberration in Microcephalin 1 (MCPH1), resulting in the creation of a new splice site at position 6621691. MCPH1 typically regulates the G2 phase of the cell cycle, influencing the differentiation of neuronal progenitor cells. Mutations in MCPH1 are known to cause premature chromosome condensation in early G2, leading cells to enter prophase prematurely. Germline mutations typically result in cell cycle arrest, reduced proliferation, and impaired neurogenesis, while somatic mutations act as tumor suppressors, leading to increased cell proliferation.

As MCPH1 mutations are generally associated with microcephaly and short stature, this case is unusual and warrants further investigation. We hypothesize that a gain-of-function germline mutation in MCPH1 may be responsible for the patient’s condition. This would be a novel mechanism for disease associated with this gene, which shows exciting applications for MAJIQ-CLIN in mechanistic studies.

## Discussion

Clinical diagnostics of rare disease using RNA-Seq is a promising avenue to improve diagnostic rates. In this study we set out to develop a tool, MAJIQ CLIN, to aid in this clinical diagnostic task, and offer comprehensive benchmarking data to assess CLIN and similar tools. A usable clinical tool for RNA-Seq should handle complex splicing events, varying numbers of aberrant genes, and different levels of variant inclusion (PSI). It should also be user-friendly, efficient, and excel at detecting both pLSVs and oLSVs while producing a concise list with the causal gene ranked highly. With these criteria in mind, we developed MAJIQ-CLIN and evaluated it alongside tools previously proposed for this task by a variety of metrics using both synthetic and real data.

In terms of efficiency, we found MAJIQ-CLIN uses slightly more memory than LeafCutterMD and less than FRASER, but runs faster than both, dramatically so in the case of LeafCutterMD. Furthermore, MAJIQ-CLIN is also able to reuse control quantifications between runs, allowing for dynamic sample addition saving additional time and resources. In order to assess accuracy, we created the first of its kind ‘realistic’ synthetic samples and evaluated MAJIQ-CLIN, FRASER, and LeafCutterMD. Each tool was evaluated for its ability to detect pLSVs and oLSVs across various splicing events and levels of variant inclusion. Across all settings, we found MAJIQ-CLIN demonstrates high precision, significantly improving detection compared to existing tools. Notably, we found LeafCutterMD was unable to detect splicing aberrations involving intron retention. We also discovered that FRASER fails on samples that suffer from a high number of splicing aberrations, where its auto-encoder corrects those as outliers. Turning to real data, we found that using a similar batch of 30 or more samples is preferable. Our results also indicate that even if the same batch is used as the control set, including confounder correction is key for a more concise list of candidate variants. Additionally, MAJIQ-CLIN successfully solved all cases from Cummings et al. [6], and the impact of confounder correction on variant ranking was minimal. These findings highlight the importance of carefully selecting controls and set size to ensure clinically meaningful results with any diagnostic tool. Finally, we applied MAJIQ-CLIN to previously undiagnosed patients from the Undiagnosed Diseases Network. This effort led to one confirmed diagnosis and several unpublished hypothesized variants still awaiting biological validation. These cases illustrate the clinical utility of MAJIQ-CLIN and its potential to improve molecular diagnosis rates.

Beyond the quantitative assessment of the various tools, we tried to highlight the importance of easily interpretable visualization for downstream analysis of aberrant splicing. For this, the VOILA visualization package helps MAJIQ-CLIN highlight and search splicing aberrations using the LSV formulation. The LSV formulation allows MAJIQ-CLIN to detect, quantify, and visualize complex splicing events involving many exons and junctions, while remaining easy to understand. In comparison, LeafCutterMD uses intron clusters to analyze splicing, which can be challenging to interpret. On the other hand, FRASER provides exon coordinates and identifies the type of aberration detected, facilitating downstream analysis.

Despite all these demonstrated strengths of MAJIQ-CLIN, both the tool itself and the study presented here suffer from several limitations. First, MAJIQ-CLIN is not able to detect the actual genomic mutations responsible for the splicing aberrations, complicating the validation of function or tracing inheritance. This is a challenging issue because variants may not always be in cis. Additionally, MAJIQ-CLIN does not provide information on allele specificity, which hinders the validation of variants in genes with specific inheritance patterns. One solution for this may be in integrating long-read sequencing to capture more distant SNPs and allow for allele specific splicing analysis. Another potential enhancement is to incorporate deep learning models, or splicing codes, that predict the effect of mutations to help in identifying casual variants [31][32][33][34]. The prioritization system in MAJIQ-CLIN is also relatively simple, focusing on pLSVs and large-magnitude changes. However, changes with a larger ΔΨ are not always more biologically significant. Although users can filter based on genes associated with Mendelian disease, MAJIQ-CLIN does not evaluate the biological properties of the variant itself. Future integration with a tool that predicts variant pathogenicity, such as Garcia et al. [35], could offer a more comprehensive prioritization metric. Our study itself also suffers from several notable limitations. Diagnosing splicing aberrations in patients is inherently challenging, leading to a scarcity of datasets with confidently identified causal genes. Consequently, we tested MAJIQ-CLIN on a limited number of datasets, with a significant portion being synthetic. Additionally, there is inherent limitation in comparing Ψ-GAP detection as done in the oLSV comparative analysis (Figure 2C). While we used GTEX expression data quantified by RSEM and then simulated by BEERS, both of which are unrelated to MAJIQ, the event definition for each junction’s Ψ value was based on MAJIQ’s LSV formulation over splice graphs. While such analysis is fine to assess MAJIQ-CLIN, the LSV based definition of the Ψ-GAP may negatively affect the performance of other algorithms not using the LSV formulation to call outliers. Finally, we did not compare MAJIQ-CLIN’s results to those of DNA-first methods. Despite these limitations, the available data and methods clearly demonstrate the usefulness and performance of MAJIQ-CLIN.

In conclusion, we developed MAJIQ-CLIN to aid the medical community as a practical and effective solution for RNA-Seq clinical diagnostics tool. With the significant performance improvements it offers, we hope clinical diagnostic labs will be able to now incorporate RNA-Seq into their analysis pipelines, ultimately enhancing molecular diagnosis rates for patients and their families.

## Data Availability

The code for MAJIQ and VOILA are available for academic/non-commercial use at majiq.biociphers.org. Licensing information for commercial use can be found at majiq.biociphers.org/commercial.php. All processed data and code to reproduce figures will be deposited on Zenodo before publication. The MAJIQ CLIN and matching VOILA updates will be added to the MAJIQ and VOILA repository upon publication. Raw GTEx data used for the analyses in this manuscript are available in dbGaP under accession phs000424. UDN data referenced in this manuscript is available in dbGaP under accession phs001232.v5.p2. The Baralle dataset is available at https://github.com/carojoquendo/RNA_splicing_and_disease.

https://www.majiq.biociphers.org

https://www.majiq.biociphers.org/commercial.php

https://github.com/carojoquendo/RNA_splicing_and_disease

## Acknowledgments

The authors would like to thank the Barash and Bhoj labs for valuable feedback, and specifically Benjamin Wales-McGrath for testing and documentation assistance. This research was supported by National Institutes of Health Grant R01 LM013437. UDN research reported in this publication was supported by the National Institute of Neurological Disorders and Stroke of the National Institutes of Health under Award Numbers U01HG007709 and U01HG007942. The content is solely the responsibility of the authors and does not necessarily represent the official views of the National Institutes of Health.

## Data and Materials Availability

The code for MAJIQ and VOILA are available for academic/non-commercial use at majiq.biociphers.org. Licensing information for commercial use can be found at ma-jiq.biociphers.org/commercial.php. All processed data and code to reproduce figures will be deposited on Zenodo before publication. The MAJIQ CLIN and matching VOILA updates will be added to the MAJIQ and VOILA repository upon publication. Raw GTEx data used for the analyses in this manuscript are available in dbGaP under accession phs000424. UDN data referenced in this manuscript is available in dbGaP under accession phs001232.v5.p2.

## Declaration of Generative AI and AI-Assisted Technologies in the Writing Process

During the preparation of this work the author(s) used ChatGPT 3.5 and 4.0 in order to improve the flow and readability of the text. After using this tool/service, the author(s) reviewed and edited the content as needed and take(s) full responsibility for the content of the publication.

## Methods

This subsection outlines the details of the MAJIQ-CLIN pipeline, followed by a description of the synthetic and biological RNA-Seq datasets used for comparison against FRASER and LeafCutterMD. We then discuss the parameters and considerations applied to each tool to ensure a fair comparison. Lastly, we explain how the evaluations presented in the Results section were designed and executed.

### 0.6 Pipeline

MAJIQ-CLIN leverages the MAJIQ framework for splicing quantification to detect RNA splicing outliers in patients with suspected Mendelian disorders using RNA-Seq. It identifies novel and significantly altered usage of junctions and retained introns by comparing the patient’s data to a user-submitted cohort of healthy controls. After processing, MAJIQ-CLIN provides clinicians with a list of outlier genes for further investigation.

This section provides a detailed overview of the pipeline. We begin with the workflow of MAJIQ, explaining how it constructs splicegraphs and LSVs and quantifies LSVs in terms of PSI using Bayesian inference. We then discuss the process of outlier detection, confounder correction using MOCCASIN, and prioritization of novel changes.

### 0.7 MAJIQ-CLIN Workflow

#### 0.7.1 Splicing Modeling and Quantification Splicegraphs

MAJIQ-CLIN is based on the MAJIQ framework for RNA splicing modeling [22][13], hence it represents splicing changes as Local Splicing Variations (LSVs). An LSV is defined as a set of splice junctions, or “edges” in a gene’s splice graph, connecting to or from a reference exon. A set of LSVs forms a splicegraph, a graphical representation of a gene’s splicing decisions, with exons as vertices and junctions and retained introns as distinct edges (Fig. 1A). This splicegraph is constructed by combining a GFF3 annotation with RNA-Seq data.

For MAJIQ-CLIN, the final set of patient LSVs for quantification is determined by combining a patient-only build group with build groups over controls. The build groups over controls include retained introns and junctions if they are found in a minimum number of control samples.

Often, the control build groups will include the patient itself. When this is the case, MAJIQ-CLIN builds a second patient-specific splicegraph explicitly excluding the patient from its component build groups. This splicegraph is used to identify retained introns, junctions, and LSVs that are structurally novel for the patient.

Additionally, users can specify a ‘case group’ instead of an individual case in the configuration. This feature is particularly useful when studying a patient with close relatives, as it excludes any familial variants from the ‘healthy control set’.

MAJIQ-CLIN can be configured to compare many patient cases each against a common collection of controls. In order to calculate the splicegraph and Ψ coverage for each case-controls comparison without redundant calculations for the shared control LSVs, MAJIQ- CLIN takes an optimized compute path. First, the splicegraph for each case-control analysis is calculated by combining two component splicegraphs, one from the case and one from all the controls. Next, PSI coverage is calculated in two passes: a first pass for the control LSVs over all samples, and a second to capture the LSVs distinct to each case-control comparison. The splicegraph operations and two-pass psi coverage workflow are generally-usable features of MAJIQ V3[23].

#### Splicing Quantifications

To quantify splicing decisions, MAJIQ calculates Percent Spliced In, a measure of junction usage, (PSI, or Ψ) for each junction in an LSV. Rather than calculate Ψ as a simple ratio of reads, MAJIQ estimates a Ψ distribution for each event. Specifically, MAJIQ reports a beta distribution for Ψ calculated from a binomial likelihood and generalized Jeffrey’s prior. The variance of the Ψ distribution reflects the level of certainty in MAJIQ’s estimate of PSI based on the number of RNA-Seq reads and PSI variation over bootstrap samples from the positions spanning the splice junctions (see MAJIQ [22] for details). MAJIQ-CLIN extracts the mean and variance of the resulting mixture of these *β* distributions, and matches a new *β* distribution to that mean and variance for a smooth approximation of the mixture of bootstrapped posteriors. For the patient sample, this smoothed distribution is used in downstream calculations. For the controls, the final distribution of Ψ for a junction is given by a distribution over the empirical means of the distributions for each sample.

### 0.7.2 Outlier Detection

#### Confounder Correction

MAJIQ-CLIN first produces uncorrected Ψ coverage for each patient and the controls to which they will be compared (which may be different than the controls used to build the splicegraph). Then, MAJIQ-CLIN allows users to correct for known confounders (eg. sequencing lane) and unknown confounders (ex. a diet change never recorded) using an integrated implementation of MOCCASIN[14]. MAJIQ-CLIN’s new MOCCASIN implementation, integrated via the new MAJIQ V3 engine, also includes updates to MOCCASIN such as vectorization over junctions and bootstrap replicates, removal of unnecessary steps, multi-threading and/or distributed parallelism. These enhancements significantly improve the efficiency of CLIN.

#### Outlier Detection using **Ψ**-GAP

As described above, MAJIQ-CLIN detects outliers of two types: pLSVs and oLSVs. pLSVs are splicing junctions unique to the patient, detected based on whether their inclusion changes the structure of an LSV. They are permitted in at most `min experiments` individuals in the control distribution (with a default value of 1). oLSVs, splicing junctions not unique but used significantly more/less in the patient, are detected by a gap between the extreme quantiles of the controls (from empirical distribution of Ψ posterior means) and the patient (from posterior distribution of Ψ) called the Ψ-GAP.

The Ψ-GAP is parameterized by a parameter *α*. *α* is used similarly to a “significance level” from two-sided null hypothesis tests. Specifically, we calculate the quantiles corresponding to 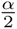 and 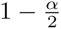 for each distribution described above. When the intervals defined by these quantiles overlap, the *ψ_gap_* is 0; otherwise, the *ψ_gap_* is quantified as the difference between the closest quantiles. That is, if *ψ*_{_*_controls,patient_*_}_(*q*) is the *q*-th quantile from the controls or patient distribution, we define

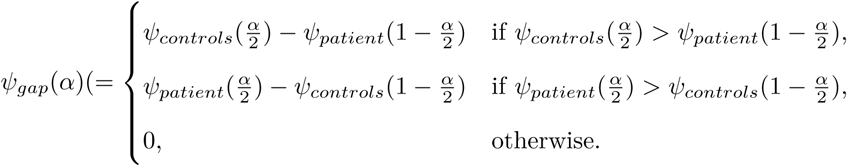

Figure 1 provides a graphical illustration.

If Ψ-GAP is sufficiently large (with a default threshold of 0.1), we identify the junction as an outlier. We only identify outliers for events which are quantifiable in the patient and over controls (quantifiable in some minimum number of experiments in controls, default 75%). We also ignore gaps where the difference in Ψ between the median of controls and patient posterior mean is less than some minimum threshold (default 10%).

It’s crucial for likely causal variants to be prioritized at the top of the list returned to the clinician. MAJIQ-CLIN lists pLSVs first, followed by oLSVs, and ranked in order of decreasing Ψ-GAP. Additionally, CLIN offers the option to filter based on a list of known protein-coding genes, known disease-causing genes from OMIM, or another user-submitted list, enabling the selection of variants of particular interest.

### 0.8 Datasets

#### 0.8.1 Simulated Datasets

We created a comprehensive synthetic dataset to evaluate MAJIQ-CLIN, LeafCutterMD, and FRASER in terms of their ability to detect oLSVs and pLSVs. This synthetic approach provided a known ground truth and a large number of samples often lacking in biological splicing datasets. It also allowed us to evaluate tool performance on different splicing aberrations and levels of junction inclusion. These synthetic datasets were generated using the BEERS [25] simulator.

##### BEERS Simulation

We used publicly-available expression quantification data from GTEx v8 as the basis for our simulations. We note that these quantifications are based on RSEM [36] and are unrelated to MAJIQ, FRASER, or LeafCutterMD. We used the GRCh38 reference genome and GENCODE v26 annotation across all simulated samples.

To simulated strand-specific reads with uniform coverage with no errors, substitutions, or intron retention events, we ran the BEERS simulator using the following command-line options: `-strandspecific -outputfq -error 0 -subfreq 0 -indelfreq 0 -intronfreq 0 -palt 0 -fraglength 100,250,500`

##### pLSV set

Four samples across two tissues (GTEX-1KWVE-2226-SM-CXZKN, GTEX-15E06-2926-SM- 6M48D, GTEX-18QFQ-0326-SM-731C3 and GTEX-1LG7Z-2126-SM-CXKYU) were selected from GTEx to serve as the basis for simulated samples. Selected samples were downloaded as FASTQ files using SRA Tools (v2.9.6). Each GTEx sample provided the gene expression distributions underlying a set of simulated samples.

Using GENCODE, we obtained a list of constitutive transcript features: junctions, exons, 5’ and 3’ splice sites. Splice sites at the start or end of transcripts, as well as overlapping genes, were excluded. We then selected genes containing at least one constitutive feature and introduced splicing aberrations across all transcripts containing it as per Table 1.

**Table 1:** Mapping between constitutive features and their corresponding splicing aberrations.

| Constitutive Feature | Splicing Aberration |
| --- | --- |
| Junction | New exon, IR |
| Exon | Exon skipping |
| 5' Splice Site | 5' exon extension/shortening |
| 3' Splice Site | 3' exon extension/shortening |

Originally, we planned to use the above data generation scheme to test all algorithms on synthetic “patient” samples that contained thousands of these aberrations such that we could get robust assessment of each algorithm performance across the various aberrations types and levels of inclusion levels. However, such a testing scheme could not be used with FRASER as its internal confounder correction flagged the “patient” sample as an outlier due to the thousands of splicing changes and corrected it.

Given the above testing limitation we resorted to the data generation scheme described next where each sample was introduced with only 5-100 aberrant genes such that FRASER will not correct it. Using this list of features and the transcript quantifications of the original GTEx samples, we generated 100 synthetic experiments. For each, we randomly chose genes for which to replace transcripts with the aberrant ones generated above, such that each sample would fulfill the coverage requirements outlined in Table 2. The aberrant transcripts were introduced at one of five levels of Ψ (0.05, 0.1, 0.2 or 0.5). We then generated coverage distributions from these transcript quantifications and sampled reads from them, all using BEERS.

**Table 2:** Minimum TPM requirements for aberrant genes in each synthetic sample.

| Level | Num. Aberrant Genes | Min. TPM Requirements |
| --- | --- | --- |
| 1 | 5 | 100 |
| 2 | 10 | 5 with min. 100, 5 with min. 10 |
| 3 | 25 | 5 with min. 100, 15 with min. 10, 5 with min. 1 |
| 4 | 50 | 10 with min. 100, 30 with min. 10, 10 with min. 1 |
| 5 | 100 | 10 with min. 100, 70 with min. 10, 20 with min. 1 |

##### oLSV set

We selected 300 samples from GTEx (150 Cerebellum, 150 Muscle-Skeletal) to create simulated samples, with each GTEx sample providing the gene expression distributions for one simulated sample. This dataset was previously used to validate MAJIQ V2 [13]. We randomly selected 20 cerebellum and 20 skeletal muscle RNA-Seq experiments to use as the evaluation “cases” for all tools, with the samples from the other tissue serving as controls.

The BEERS simulation output provided transcript-level TPM quantifications for each sample, serving as the TPM ground truth. We used MAJIQ V3 to define the LSVs in the data and then quantified the ground truth for those LSVs and associated splice junctions using the aforementioned BEERS TPMs. These values then served as ground truths to compare Ψ-GAP (*G*) detection at one of three thresholds (*G* ∈ {0.1, 0.2, 0.3}).

The above oLSV data generation enabled us to reuse previously generated data efficiently and create “natural” splice variations in terms of both coverage and occurrence across human donor tissues. However, given the many hundreds of splicing variations per “case” created this way we were unable to test FRASER on this data, as its algorithm seems to automatically “corrects” the case sample.

#### 0.8.2 Patient Datasets

We analyzed patient RNA-Seq samples from three different datasets: (1) the Cummings dataset [6], (2) the Baralle dataset [21], and (3) the UDN dataset [20]. The Cummings and UDN datasets were downloaded from dbGaP, while the Baralle dataset is available at https://github.com/carojoquendo/RNA_splicing_and_disease.

The Cummings dataset is composed of 53 RNA-Seq samples from muscle biopsies of patients with previously genetically undiagnosed rare muscle disorders. From these data and matched exome or genome sequencing, Cummings et al. (2017) identified molecular diagnoses in 25 patients. Of these, we determined that 16 patients were candidates for RNA-first detection as splicing outliers (the others: 4 allele-specific expression, 2 large structural variants, and 3 core-splice site variants identified from genetic sequencing but with insufficient coverage for detection from RNA-Seq). All samples were sequenced at the same site, so we treated these data as if there were no confounders.

The Baralle dataset is composed of 55 RNA-Seq samples from globin-depleted whole blood from patients with suspected Mendelian disorders with diverse phenotypes. These data were sequenced in three batches with 7, 16, and 32 samples each, so we used batch identity as a confounder in our analyses.

The UDN dataset is composed of 400 RNA-Seq samples from fibroblasts from patients with suspected Mendelian disorders with diverse phenotypes and their affected/unaffected relatives. These data were sequenced with four different sequencing instruments, so we included sequencing instrument as a known confounder.

While batch correction and detection of outliers were limited to samples from within each individual dataset, we included samples with RIN score greater than 6 from GTEx v8 as additional build groups for constructing splicegraphs. For the Cummings dataset, we included all skeletal muscle samples as a build group. For the Baralle dataset, we included all GTEx whole blood and EBV-transformed lymphocytes samples as build groups. For the UDN dataset, we included all GTEx cultured fibroblasts as a build group.

### 0.9 Evaluations

#### 0.9.1 Pre-processing

All data was originally in the FASTQ format. We performed quality and adapter trimming on each sample using TrimGalore (v0.4.5). We used STAR (v2.5.3a) to perform a two-step gapped alignment of the trimmed reads to the GRCh38 primary assembly with annotations from Ensembl release 94.

#### 0.9.2 Tool Setup

##### LeafCutterMD

LeafCutterMD was run with default parameters. We removed any filtering of results based on `padj_threshold` so that we could manually adjust `padj_threshold` when analyzing results. Unadjusted (raw) p-values were used for ROC/PR curves, as lossy p-value adjustment can push insignificant p-values to 1.

For analyses excluding runtime and efficiency, some adjustments were made to the LeafCutterMD pipeline to speed up the pre-analysis steps. By default, the LeafCutterMD provided “bam2junc” program does not allow parallel processing, overwriting junc files in the event of two runs using the same bam file. We overcame this bottleneck by setting up a Snakemake pipeline to run LeafCutterMD and replacing the default bam2junc program with one which used junc files from a cached location. We also added another program to collectively evaluate all required bamfiles for all experiments prior to starting a run of LeafCutterMD.

##### FRASER

For FRASER the FRASER2 [19] version was used for all evaluations. While no further versions have been released, the FRASER2 GitHub has been updated with bug fixes. All analyses use the latest version of the software available at the time of running.

FRASER was run with default parameters. We encountered an issue with the final step in the FRASER pipeline, where the results are converted to HTML output. As such, our FRASER runs complete at 95% without the html generation (we considered “completeness” being the generation of results_gene_all.tsv). This is the case in the runtime and memory analysis.

##### MAJIQ-CLIN

We used GENCODE V26 with the Ensembl.94.GRCh38 annotation. Each run had `controls_min_experiments` set to 1 and `case_min_experiments` set to 0.75. The build flag –`simplify` was used, activating the MAJIQ simplifier [13]. *α* was set to 0.1. Unless noted otherwise, no confounders were modeled with MOCCASIN, and leave-one-out controls were not used.

#### 0.9.3 Analysis

##### Runtime and Efficiency

We compared MAJIQ-CLIN to its competitors on wall-clock run time and memory usage. We ran each tool on the same subsets of samples from the pLSV set, in groups of 10, 20, 30, etc., on the Children’s Hospital of Philadelphia High Performance Cluster. Runtime and memory use were reported by SLURM [37], starting each time with fresh BAM input files. It is notable that as these tests were run on a shared cluster environment, available resources may have varied slightly, which accounts for some variance. One outlier point (1 threads, 50 samples) was removed from the FRASER runs due to a run error.

Both LeafCutterMD and MAJIQ-CLIN convert BAM files to pipeline-specific intermediates for downstream processing (bam-to-junc for LeafCutterMD, bam-to-sj for MAJIQ-CLIN). These steps only need to be run once for each sample. LeafCutterMD takes an average of 37 minutes and 42 seconds to convert a BAM to a junc file, and MAJIQ-CLIN takes an average of 5 minutes 33 seconds to convert a BAM to an intermediate SJ file. For MAJIQ-CLIN, this is part of the pipeline, and done in parallel; for LeafCutterMD, the user must run an external script. LeafCutterMD run times are therefore estimated by adding the time for the bam-to-junc conversion of the files (extrapolated from an average runtime) to the runtime of the steps of LeafCutterMD post- `bamtojunc`. A Snakemake wrapper was used for LeafCutterMD functions to allow batch runs.

##### pLSV Analysis

We compared MAJIQ-CLIN, LeafCutterMD, and FRASER on the pLSV set, with oLSV samples from appropriate tissues serving as controls. Samples derived from the same original GTEx data were excluded from the control set to avoid bias. Sensitivity, specificity, and precision were quantified using the set of transcripts initially defined for BEERS as the ground truth. Results were aggregated at the gene level to ensure an equivalent comparison across tools. Specifically, a gene was considered an expected positive if it had an aberrant transcript added in a sample and an expected negative if it only retained its original transcripts.

In the MAJIQ-CLIN results, a gene was considered a positive hit if any junction within it had a quantile gap of at least the quantile gap threshold (varying from -1 to 1) and had at least 10 reads, and a negative otherwise. For LeafCutterMD, a gene was considered a positive hit if any intron cluster within the gene was identified as an outlier with a p-value less than the p-value threshold (varying 1 from to 1 exp −20), as well as having an absolute dPSI of at least 0.1, and a negative otherwise. For FRASER, a gene was considered a positive hit if it had a dPSI of at least 0.1 and a p-value less than the p-value threshold (varying from 1 to 1 exp −20) and negative otherwise.

##### oLSV analysis

To evaluate the ability of the tools to detect oLSVs, we took advantage of the natural splicing variation between tissues. We randomly selected 20 Cerebellum and 20 Skeletal Muscle RNA-Seq experiments to use as “cases” from the oLSV set described above. We then executed MAJIQ-CLIN in one run containing two analyses, one with the 150 cerebellum samples each run against 150 muscle skeletal controls, and the second with the tissues swapped. (The results for only 20 cases for each tissue were ultimately used.)

To establish the ground truth outliers for this analysis, we ran MAJIQ V2 with the 20 ‘cases’ and 150 ‘controls’ for each tissue. To constrain the evaluation to oLSVs, only LSVs quantified in the case and at least one control were considered. For each of these shared LSVs, the 5th (p5) and 95th (p95) percentile E[Ψ] values were calculated across all control samples. The Ψ*_GAP_* is defined as the greater of (p5 - E[Ψ]) and (E[Ψ] - p95). This captures any arrangement of the case and control Ψ distributions.

Ψ*_GAP_* threshold *G* is the threshold that an LSV must pass to be called as an outlier by MAJIQ-CLIN. In the subsequent analysis, this threshold was varied from 0 to 1. For a fixed value of Ψ*_GAP_* threshold *G*^∗^, an LSV is a “ground truth outlier” if at least one of its junctions has Ψ*_GAP_ > G*^∗^. This compares the mean of the ground-truth Ψ distribution to the control quantiles, as compared with MAJIQ-CLIN, which compares the extreme quantiles for the case distribution to the control quantiles as described above. A gene is an expected positive if it contains at least one “ground truth outlier” LSV.

As before, a gene was considered a positive hit for LeafCutterMD if any intron cluster was identified as an outlier with a p-value below a threshold varied from 1 to 10^−20^. For MAJIQ-CLIN, outliers are also called similarly to the pLSV analysis. A gene is considered a positive if any LSV within it has a Ψ*_GAP_* greater than *G*^∗^ threshold, ranging from 0 to 1, with the pLSV flag not being considered. FRASER was not included in this evaluation due to breaking changes in its code and dependencies that prevented it from running successfully at the time.

##### Confounder Correction and Ranking

Controls were used for the Baralle samples as described in Results, with default settings. The GTEx Muscle-Skeletal set was used as the control set for the Cummings samples. Rank was determined by first occurrence in the list of a gene whose HGNC ID matched the solved gene from Cummings et al. Two samples had two runs associated with them, C1 and N32. SRR5034830 was used for C1, and SRR5020918 was used for N32. No known confounders were specified.

## References

[1] Stéphanie Nguengang Wakap, Deborah M. Lambert, Annie Olry, Charlotte Rodwell, Charlotte Gueydan, Valérie Lanneau, Daniel Murphy, Yann Le Cam, and Ana Rath. Estimating cumulative point prevalence of rare diseases: analysis of the Orphanet database. European Journal of Human Genetics, 28(2):165–173, February 2020. Publisher: Nature Publishing Group.

[2] Caroline F. Wright, David R. FitzPatrick, and Helen V. Firth. Paediatric genomics: diagnosing rare disease in children. Nature Reviews. Genetics, 19(5):253–268, May 2018.

[3] McKusick-Nathans Institute of Genetic Medicine. Online mendelian inheritance in man, OMIM®, Accessed 2024-10-03. World Wide Web URL: https://omim.org/.

[4] Anne Slavotinek, Shannon Rego, Nuriye Sahin-Hodoglugil, Mark Kvale, Billie Lianoglou, Tiffany Yip, Hannah Hoban, Simon Outram, Beatrice Anguiano, Flavia Chen, Jeremy Michelson, Roberta M. Cilio, Cynthia Curry, Renata C. Gallagher, Marisa Gardner, Rachel Kuperman, Bryce Mendelsohn, Elliott Sherr, Joseph Shieh, Jonathan Strober, Allison Tam, Jessica Tenney, William Weiss, Amy Whittle, Garrett Chin, Amanda Faubel, Hannah Prasad, Yusuph Mavura, Jessica Van Ziffle, W. Patrick Devine, Ugur Hodoglugil, Pierre-Marie Martin, Teresa N. Sparks, Barbara Koenig, Sara Ackerman, Neil Risch, Pui-Yan Kwok, and Mary E. Norton. Diagnostic yield of pediatric and prenatal exome sequencing in a diverse population. npj Genomic Medicine, 8(1):1–10, May 2023. Publisher: Nature Publishing Group.

[5] Zornitza Stark, Tiong Y. Tan, Belinda Chong, Gemma R. Brett, Patrick Yap, Maie Walsh, Alison Yeung, Heidi Peters, Dylan Mordaunt, Shannon Cowie, David J. Amor, Ravi Savarirayan, George McGillivray, Lilian Downie, Paul G. Ekert, Christiane Theda, Paul A. James, Joy Yaplito-Lee, Monique M. Ryan, Richard J. Leventer, Emma Creed, Ivan Macciocca, Katrina M. Bell, Alicia Oshlack, Simon Sadedin, Peter Georgeson, Charlotte Anderson, Natalie Thorne, null Melbourne Genomics Health Alliance, Clara Gaff, and Susan M. White. A prospective evaluation of whole-exome sequencing as a first-tier molecular test in infants with suspected monogenic disorders. Genetics in Medicine: Official Journal of the American College of Medical Genetics, 18(11):1090– 1096, November 2016.

[6] B. B. Cummings, J. L. Marshall, T. Tukiainen, M. Lek, S. Donkervoort, A. R. Foley, V. Bolduc, L. B. Waddell, S. A. Sandaradura, G. L. O’Grady, E. Estrella, H. M. Reddy, F. Zhao, B. Weisburd, K. J. Karczewski, A. H. O’Donnell-Luria, D. Birnbaum, A. Sarkozy, Y. Hu, H. Gonorazky, K. Claeys, H. Joshi, A. Bournazos, E. C. Oates, R. Ghaoui, M. R. Davis, N. G. Laing, A. Topf, Genotype-Tissue Expression Consortium, P. B. Kang, A. H. Beggs, K. N. North, V. Straub, J. J. Dowling, F. Muntoni, N. F. Clarke, S. T. Cooper, C. G. Bönnemann, and D. G. MacArthur. Improving genetic diagnosis in mendelian disease with transcriptome sequencing. Science Translational Medicine, 9(386):eaal5209, April 2017.

[7] Joris Deelen, Daniel S. Evans, Dan E. Arking, Niccolò Tesi, Marianne Nygaard, Xiaomin Liu, Mary K. Wojczynski, Mary L. Biggs, Ashley van der Spek, Gil Atzmon, Erin B. Ware, Chloé Sarnowski, Albert V. Smith, Ilkka Seppälä, Heather J. Cordell, Janina Dose, Najaf Amin, Alice M. Arnold, Kristin L. Ayers, Nir Barzilai, Elizabeth J. Becker, Marian Beekman, Hélène Blanché, Kaare Christensen, Lene Christiansen, Joanna C. Collerton, Sarah Cubaynes, Steven R. Cummings, Karen Davies, Birgit Debrabant, Jean-François Deleuze, Rachel Duncan, Jessica D. Faul, Claudio Franceschi, Pilar Galan, Vilmundur Gudnason, Tamara B. Harris, Martijn Huisman, Mikko A. Hurme, Carol Jagger, Iris Jansen, Marja Jylhä, Mika Kähönen, David Karasik, Sharon L. R. Kardia, Andrew Kingston, Thomas B. L. Kirkwood, Lenore J. Launer, Terho Lehtimäki, Wolfgang Lieb, Leo-Pekka Lyytikäinen, Carmen Martin-Ruiz, Junxia Min, Almut Nebel, Anne B. Newman, Chao Nie, Ellen A. Nohr, Eric S. Orwoll, Thomas T. Perls, Michael A. Province, Bruce M. Psaty, Olli T. Raitakari, Marcel J. T. Reinders, Jean-Marie Robine, Jerome I. Rotter, Paola Sebastiani, Jennifer Smith, Thorkild I. A. Sørensen, Kent D. Taylor, André G. Uitterlinden, Wiesje van der Flier, Sven J. van der Lee, Cornelia M. van Duijn, Diana van Heemst, James W. Vaupel, David Weir, Kenny Ye, Yi Zeng, Wanlin Zheng, Henne Holstege, Douglas P. Kiel, Kathryn L. Lunetta, P. Eline Slagboom, and Joanne M. Murabito. A meta-analysis of genome-wide association studies identifies multiple longevity genes. Nature Communications, 10(1):3669, August 2019. Publisher: Nature Publishing Group.

[8] Ashish R. Deshwar, Kyoko E. Yuki, Huayun Hou, Yijing Liang, Tayyaba Khan, Alper Celik, Arun Ramani, Roberto Mendoza-Londono, Christian R. Marshall, Michael Brudno, Adam Shlien, M. Stephen Meyn, Robin Z. Hayeems, Brandon J. McKinlay, Panagiota Klentrou, Michael D. Wilson, Lianna Kyriakopoulou, Gregory Costain, and James J. Dowling. Trio RNA sequencing in a cohort of medically complex children. American Journal of Human Genetics, 110(5):895–900, May 2023.

[9] Kevin Riquin, Bertrand Isidor, Sandra Mercier, Mathilde Nizon, Estelle Colin, Dominique Bonneau, Laurent Pasquier, Sylvie Odent, Xavier Maximin Le Guillou Horn, Gwenaël Le Guyader, Annick Toutain, Vincent Meyer, Jean-François Deleuze, Olivier Pichon, Martine Doco-Fenzy, Stéphane Bézieau, and Benjamin Cogné. Integrating RNA-Seq into genome sequencing workflow enhances the analysis of structural variants causing neurodevelopmental disorders. Journal of Medical Genetics, 61(1):47–56, December 2023.

[10] Qun Pan, Ofer Shai, Leo J. Lee, Brendan J. Frey, and Benjamin J. Blencowe. Deep surveying of alternative splicing complexity in the human transcriptome by high-throughput sequencing. Nature Genetics, 40(12):1413–1415, December 2008. Publisher: Nature Publishing Group.

[11] Lea M. Starita, Nadav Ahituv, Maitreya J. Dunham, Jacob O. Kitzman, Frederick P. Roth, Georg Seelig, Jay Shendure, and Douglas M. Fowler. Variant Interpretation: Functional Assays to the Rescue. American Journal of Human Genetics, 101(3):315–325, September 2017.

[12] Joseph K. Aicher, Paul Jewell, Jorge Vaquero-Garcia, Yoseph Barash, and Elizabeth J. Bhoj. Mapping RNA splicing variations in clinically accessible and nonaccessible tissues to facilitate Mendelian disease diagnosis using RNA-seq. Genetics in Medicine: Official Journal of the American College of Medical Genetics, 22(7):1181–1190, July 2020.

[13] Jorge Vaquero-Garcia, Joseph K. Aicher, San Jewell, Matthew R. Gazzara, Caleb M. Radens, Anupama Jha, Scott S. Norton, Nicholas F. Lahens, Gregory R. Grant, and Yoseph Barash. RNA splicing analysis using heterogeneous and large RNA-seq datasets. Nature Communications, 14(1):1230, March 2023.

[14] Barry Slaff, Caleb M. Radens, Paul Jewell, Anupama Jha, Nicholas F. Lahens, Gregory R. Grant, Andrei Thomas-Tikhonenko, Kristen W. Lynch, and Yoseph Barash. MOCCASIN: a method for correcting for known and unknown confounders in RNA splicing analysis. Nature Communications, 12(1):3353, June 2021. Publisher: Nature Publishing Group.

[15] Hernan D. Gonorazky, Sergey Naumenko, Arun K. Ramani, Viswateja Nelakuditi, Pouria Mashouri, Peiqui Wang, Dennis Kao, Krish Ohri, Senthuri Viththiyapaskaran, Mark A. Tarnopolsky, Katherine D. Mathews, Steven A. Moore, Andres N. Osorio, David Villanova, Dwi U. Kemaladewi, Ronald D. Cohn, Michael Brudno, and James J. Dowling. Expanding the Boundaries of RNA Sequencing as a Diagnostic Tool for Rare Mendelian Disease. American Journal of Human Genetics, 104(3):466–483, March 2019.

[16] Garrett Jenkinson, Yang I Li, Shubham Basu, Margot A Cousin, Gavin R Oliver, and Eric W Klee. LeafCutterMD: an algorithm for outlier splicing detection in rare diseases. Bioinformatics, 36(17):4609–4615, November 2020.

[17] Yang I. Li, David A. Knowles, Jack Humphrey, Alvaro N. Barbeira, Scott P. Dickinson, Hae Kyung Im, and Jonathan K. Pritchard. Annotation-free quantification of RNA splicing using LeafCutter. Nature Genetics, 50(1):151–158, January 2018.

[18] Christian Mertes, Ines F. Scheller, Vicente A. Yépez, Muhammed H. Çelik, Yingjiqiong Liang, Laura S. Kremer, Mirjana Gusic, Holger Prokisch, and Julien Gagneur. Detection of aberrant splicing events in RNA-seq data using FRASER. Nature Communications, 12(1):529, January 2021. Publisher: Nature Publishing Group.

[19] Ines F. Scheller, Karoline Lutz, Christian Mertes, Vicente A. Yépez, and Julien Gagneur. Improved detection of aberrant splicing with FRASER 2.0 and the intron Jaccard index. The American Journal of Human Genetics, 110(12):2056–2067, December 2023.

[20] David R. Murdock, Hongzheng Dai, Lindsay C. Burrage, Jill A. Rosenfeld, Shamika Ketkar, Michaela F. Müller, Vicente A. Yépez, Julien Gagneur, Pengfei Liu, Shan Chen, Mahim Jain, Gladys Zapata, Carlos A. Bacino, Hsiao-Tuan Chao, Paolo Moretti, William J. Craigen, Neil A. Hanchard, and Brendan Lee. Transcriptome-directed analysis for Mendelian disease diagnosis overcomes limitations of conventional genomic testing. The Journal of Clinical Investigation, 131(1), January 2021. Publisher: American Society for Clinical Investigation.

[21] Carolina Jaramillo Oquendo, Htoo A. Wai, Wil I. Rich, David J. Bunyan, N. Simon Thomas, David Hunt, Jenny Lord, Andrew G. L. Douglas, and Diana Baralle. Identification of diagnostic candidates in Mendelian disorders using an RNA sequencing-centric approach. Genome Medicine, 16(1):110, September 2024.

[22] Jorge Vaquero-Garcia, Alejandro Barrera, Matthew R Gazzara, Juan González-Vallinas, Nicholas F Lahens, John B Hogenesch, Kristen W Lynch, and Yoseph Barash. A new view of transcriptome complexity and regulation through the lens of local splicing variations. eLife, 5:e11752, February 2016. Publisher: eLife Sciences Publications, Ltd.

[23] Joseph K. Aicher, Barry Slaff, San Jewell, and Yoseph Barash. MAJIQ V3 offers improvements in accuracy, performance, and usability for splicing analysis from RNA sequencing, July 2024. Pages: 2024.07.02.601792 Section: New Results.

[24] Alexander J. M. Blakes, Htoo A. Wai, Ian Davies, Hassan E. Moledina, April Ruiz, Tessy Thomas, David Bunyan, N. Simon Thomas, Christine P. Burren, Lynn Greenhalgh, Melissa Lees, Amanda Pichini, Sarah F. Smithson, Ana Lisa Taylor Tavares, Peter O’Donovan, Andrew G. L. Douglas, Nicola Whiffin, Diana Baralle, Jenny Lord, and Splicing and Disease Working Group Genomics England Research Consortium. A systematic analysis of splicing variants identifies new diagnoses in the 100,000 Genomes Project. Genome Medicine, 14(1):79, July 2022.

[25] Gregory R. Grant, Michael H. Farkas, Angel D. Pizarro, Nicholas F. Lahens, Jonathan Schug, Brian P. Brunk, Christian J. Stoeckert, John B. Hogenesch, and Eric A. Pierce. Comparative analysis of RNA-Seq alignment algorithms and the RNA-Seq unified mapper (RUM). Bioinformatics, 27(18):2518–2528, September 2011.

[26] Ying Ge and Bo T. Porse. The functional consequences of intron retention: alternative splicing coupled to NMD as a regulator of gene expression. *BioEssays: News and Reviews in Molecular*, Cellular and Developmental Biology, 36(3):236–243, March 2014.

[27] Ulrich Braunschweig, Nuno L. Barbosa-Morais, Qun Pan, Emil N. Nachman, Babak Alipanahi, Thomas Gonatopoulos-Pournatzis, Brendan Frey, Manuel Irimia, and Benjamin J. Blencowe. Widespread intron retention in mammals functionally tunes transcriptomes. Genome Research, 24(11):1774–1786, November 2014.

[28] Heidi Dvinge and Robert K. Bradley. Widespread intron retention diversifies most cancer transcriptomes. Genome Medicine, 7(1):45, 2015.

[29] Hong-Dong Li, Cory C. Funk, Karen McFarland, Eric B. Dammer, Mariet Allen, Minerva M. Carrasquillo, Yona Levites, Paramita Chakrabarty, Jeremy D. Burgess, Xue Wang, Dennis Dickson, Nicholas T. Seyfried, Duc M. Duong, James J. Lah, Steven G. Younkin, Allan I. Levey, Gilbert S. Omenn, Nilüfer Ertekin-Taner, Todd E. Golde, and Nathan D. Price. Integrative functional genomic analysis of intron retention in human and mouse brain with Alzheimer’s disease. Alzheimer’s & Dementia, 17(6):984–1004, June 2021.

[30] Hirotomo Saitsu, Hitoshi Osaka, Masayuki Sasaki, Jun-ichi Takanashi, Keisuke Hamada, Akio Yamashita, Hidehiro Shibayama, Masaaki Shiina, Yukiko Kondo, Kiyomi Nishiyama, Yoshinori Tsurusaki, Noriko Miyake, Hiroshi Doi, Kazuhiro Ogata, Ken Inoue, and Naomichi Matsumoto. Mutations in POLR3A and POLR3B Encoding RNA Polymerase III Subunits Cause an Autosomal-Recessive Hypomyelinating Leukoencephalopathy. American Journal of Human Genetics, 89(5):644–651, November 2011.

[31] Pedro Barbosa, Rosina Savisaar, Maria Carmo-Fonseca, and Alcides Fonseca. Computational prediction of human deep intronic variation. GigaScience, 12:giad085, January 2023.

[32] Jun Cheng, Thi Yen Duong Nguyen, Kamil J. Cygan, Muhammed Hasan Çelik, William G. Fairbrother, žiga Avsec, and Julien Gagneur. MMSplice: modular modeling improves the predictions of genetic variant effects on splicing. Genome Biology, 20(1):48, March 2019.

[33] Kishore Jaganathan, Sofia Kyriazopoulou Panagiotopoulou, Jeremy F. McRae, Siavash Fazel Darbandi, David Knowles, Yang I. Li, Jack A. Kosmicki, Juan Arbelaez, Wenwu Cui, Grace B. Schwartz, Eric D. Chow, Efstathios Kanterakis, Hong Gao, Amirali Kia, Serafim Batzoglou, Stephan J. Sanders, and Kyle Kai-How Farh. Predicting Splicing from Primary Sequence with Deep Learning. Cell, 176(3):535–548.e24, January 2019.

[34] Hui Y. Xiong, Babak Alipanahi, Leo J. Lee, Hannes Bretschneider, Daniele Merico, Ryan K. C. Yuen, Yimin Hua, Serge Gueroussov, Hamed S. Najafabadi, Timothy R. Hughes, Quaid Morris, Yoseph Barash, Adrian R. Krainer, Nebojsa Jojic, Stephen W. Scherer, Benjamin J. Blencowe, and Brendan J. Frey. The human splicing code reveals new insights into the genetic determinants of disease. Science, 347(6218):1254806, January 2015. Publisher: American Association for the Advancement of Science.

[35] Felipe Antonio de Oliveira Garcia, Edilene Santos de Andrade, and Edenir Inez Palmero. Insights on variant analysis in silico tools for pathogenicity prediction. Frontiers in Genetics, 13:1010327, November 2022.

[36] Bo Li and Colin N. Dewey. RSEM: accurate transcript quantification from RNA-Seq data with or without a reference genome. BMC Bioinformatics, 12(1):323, August 2011.

[37] Andy B. Yoo, Morris A. Jette, and Mark Grondona. Slurm: Simple linux utility for resource management. In Dror Feitelson, Larry Rudolph, and Uwe Schwiegelshohn, editors, Job Scheduling Strategies for Parallel Processing, pages 44–60, Berlin, Heidelberg, 2003. Springer Berlin Heidelberg.

